# Rapidly shifting immunologic landscape and severity of SARS-CoV-2 in the Omicron era in South Africa

**DOI:** 10.1101/2022.08.19.22278993

**Authors:** Kaiyuan Sun, Stefano Tempia, Jackie Kleynhans, Anne von Gottberg, Meredith L McMorrow, Nicole Wolter, Jinal N. Bhiman, Jocelyn Moyes, Maimuna Carrim, Neil A Martinson, Kathleen Kahn, Limakatso Lebina, Jacques D. du Toit, Thulisa Mkhencele, Cécile Viboud, Cheryl Cohen, the PHIRST group

## Abstract

South Africa was among the first countries to detect the SARS-CoV-2 Omicron variant. Propelled by increased transmissibility and immune escape properties, Omicron displaced other globally circulating variants within 3 months of its emergence. Due to limited testing, Omicron’s attenuated clinical severity, and an increased risk of reinfection, the size of the Omicron BA.1 and BA.2 subvariants (BA.1/2) wave remains poorly understood in South Africa and in many other countries. Using South African data from urban and rural cohorts closely monitored since the beginning of the pandemic, we analyzed sequential serum samples collected before, during, and after the Omicron BA.1/2 wave to infer infection rates and monitor changes in the immune histories of participants over time. Omicron BA.1/2 infection attack rates reached 65% (95% CI, 60% – 69%) in the rural cohort and 58% (95% CI, 61% – 74%) in the urban cohort, with repeat infections and vaccine breakthroughs accounting for >60% of all infections at both sites. Combined with previously collected data on pre-Omicron variant infections within the same cohorts, we identified 14 distinct categories of SARS-CoV-2 antigen exposure histories in the aftermath of the Omicron BA.1/2 wave, indicating a particularly fragmented immunologic landscape. Few individuals (<6%) remained naïve to SARS-CoV-2 and no exposure history category represented over 25% of the population at either cohort site. Further, cohort participants were more than twice as likely to get infected during the Omicron BA.1/2 wave, compared to the Delta wave. Prior infection with the ancestral strain (with D614G mutation), Beta, and Delta variants provided 13% (95% CI, -21% – 37%), 34% (95% CI, 17% – 48%), and 51% (95% CI, 39% – 60%) protection against Omicron BA.1/2 infection, respectively. Hybrid immunity (prior infection and vaccination) and repeated prior infections (without vaccination) reduced the risks of Omicron BA.1/2 infection by 60% (95% CI, 42% – 72%) and 85% (95% CI, 76% – 92%) respectively. Reinfections and vaccine breakthroughs had 41% (95% CI, 26% – 53%) lower risk of onward transmission than primary infections. Our study sheds light on a rapidly shifting landscape of population immunity, along with the changing characteristics of SARS-CoV-2, and how these factors interact to shape the success of emerging variants. Our findings are especially relevant to populations similar to South Africa with low SARS-CoV-2 vaccine coverage and a dominant contribution of immunity from prior infection. Looking forward, the study provides context for anticipating the long-term circulation of SARS-CoV-2 in populations no longer naïve to the virus.

## Introduction

One of SARS-CoV-2’s most prominent features has been its rapid adaptive evolution throughout the pandemic: every few months, new variants with selective advantages have emerged, displaced resident variants, and reached global dominance. To date, the World Health Organization has classified five SARS-CoV-2 lineages as variants of concern (VOCs) due to their enhanced transmissibility and immune escape properties, including Alpha, Beta, Gamma, Delta and Omicron (*1, 2*). Following the rise of Omicron (BA.1), new lineages continue to evolve with further mutations, including those on the spike protein not seen on Omicron BA.1 that evade immune responses (notably BA.2, BA.2.12.1, BA.4, and BA.5 (*3*)). The selective advantage of a new immune-escape variant and subvariants is shaped in part by the host population immunity, first at the location of emergence, and then globally (*4*).

As of August 1^st^, 2022, South Africa has experienced five SARS-CoV-2 epidemic waves: the 1^st^ wave was dominated by the ancestral strain carrying the D614G mutation (D614G); the 2^nd^ wave by the Beta VOC (with little impact of the Alpha VOC that was globally dominant at that time (*5*)); the 3^rd^ wave by the Delta VOC; the 4^th^ wave by Omicron subvariants BA.1 and BA.2 (BA.1/2 wave); and the 5^th^ wave by Omicron subvariants BA.4 and BA.5 (BA.4/5 wave). In addition to the Beta VOC (*6*), the Omicron subvariants BA.1, BA.4 and BA.5, are likely to have emerged in South Africa or the surrounding region (*7, 8*). Detailed studies of the immunologic landscape in South Africa could provide a unique perspective on how immunity contributes to variant success on a population level, near the region of their emergence. The PHIRST-C cohorts have generated detailed prospective data on infection and serology spanning South Africa’s first four waves, in carefully sampled populations from randomly selected households (*9*). Here we quantify changes in the population immunologic landscape to SARS-CoV-2 over time, and particularly in the aftermath of the Omicron BA.1/2 wave, compare Omicron’s epidemiologic properties to those of prior variants, adjusting for changes in prior immunity, and discuss how these factors may interact to determine the fate of new variants.

## Results

### Overview of serologic specimen collection and SARS-CoV-2 dynamics in the PHIRST-C cohort

As previously described (*9, 10*), in June 2020, the **P**rospective **H**ousehold study of SARS-CoV-2, **I**nfluenza and **R**espiratory **S**yncytial virus community burden, **T**ransmission dynamics and viral interaction in South Africa, **PHIRST-C**, where “**C**” stands for coronavirus disease 2019 (COVID-19) enrolled a total of 1,200 individuals living in 222 randomly selected households. Two sites were selected with 114 households (643 individuals) enrolled in a rural site located in Agincourt, Mpumalanga Province, northeast South Africa and 108 households (557 individuals) in an urban site, located in Jouberton township, Matlosana, North West Province, South Africa. From July 2020 through April 2022, ten sequential serum specimens were collected for each participant (Figure 1). Blood draws 1-9 were conducted at approximately 2-month intervals, with blood draws 8 and 9 collected one month prior and after the emergence of Omicron BA.1 variant in Southern Africa (*8*). Blood draw 10 was conducted at the end of the Omicron BA.1/2 wave, ∼4 months after blood draw 9, but prior to the emergence of Omicron BA.4/5 (Figure 1) (*8*). The cohort included a period of intense follow-up of active infections (July 16, 2020 - August 28, 2021, in the rural site and July 27, 2020 - August 28, 2021, in the urban site, covering the D614G, Beta, and Delta waves) during which nasal swab samples were collected twice-weekly for SARS-CoV-2 real-time reverse transcription polymerase chain reaction (rRT-PCR), irrespective of symptoms.

**Fig. 1.**
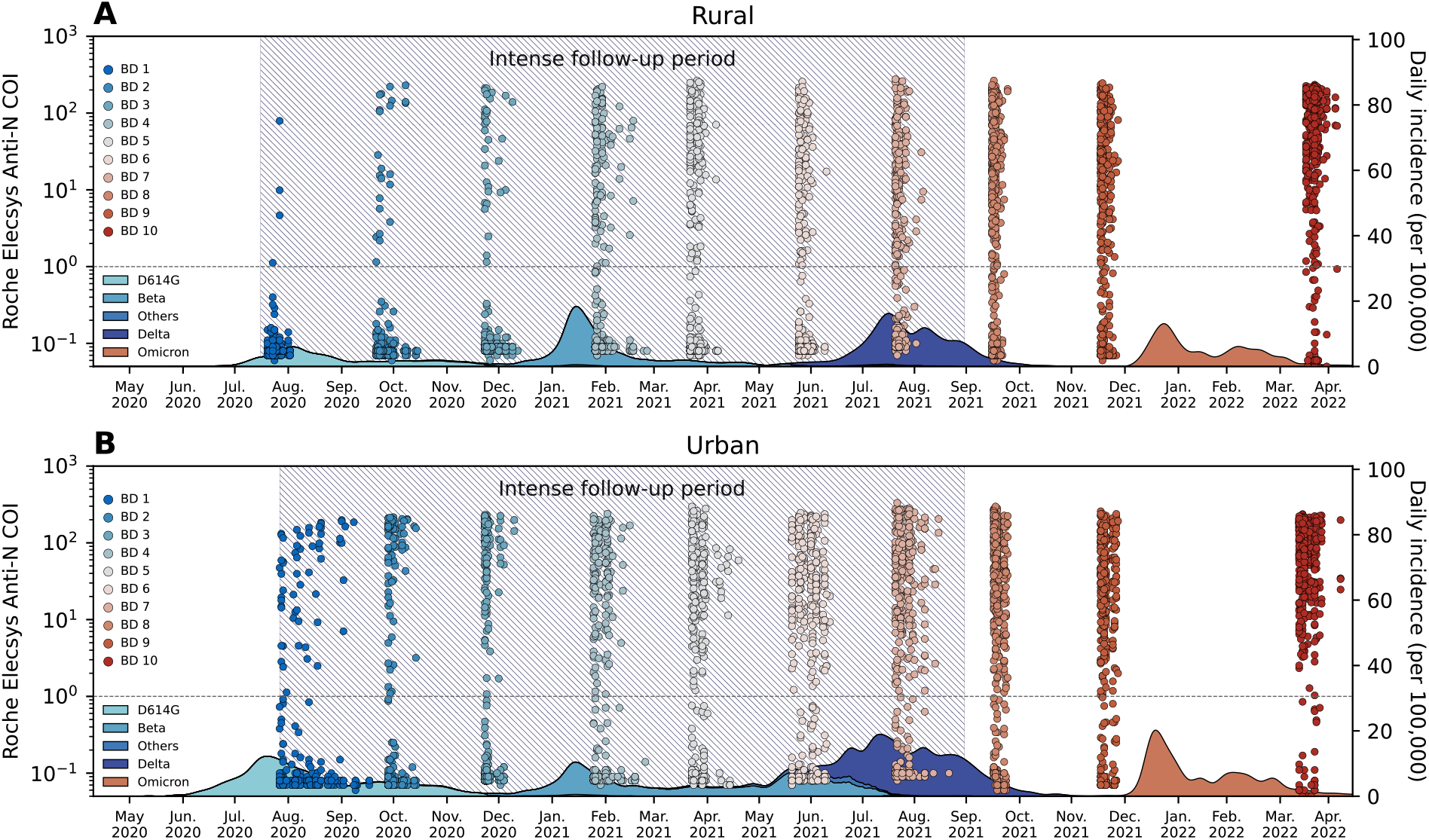
PHIRST-C study June 2020 – April 2022, SARS-CoV-2 serology and epidemiologic curve in the two study sites. (A) Serum samples and epidemiologic curve in the rural site. Dots represent the Roche Elecsys Anti-SARS-CoV-2 nucleocapsid assay cutoff index (COI) at different timepoints of the serum specimen collection; Each dot represents one serum specimen collection, with dot color denoting blood draw collection time, from blue (early) to red (late). The shaded curve at the bottom represents the daily incidence of SARS-CoV-2 cases in routine surveillance data collected by the Ehlanzeni District, Mpumalanga Province. Colors of the shaded curve represent different variant types. Here, blood draw (BD) 10 was collected at the end of the first Omicron wave. Since in South Africa, Omicron BA.4 and BA.5 only started to rise at April, 2022 (*8*), we assume the Omicron wave prior to BD 10 were BA.1 and BA.2 subvariants. The hatched area represents the period of intense follow-up of the PHIRST-C cohort, when nasal swabs were collected and tested on rRT-PCR at twice-a-week frequency. (B) Same as (A) but for the urban site. Abbreviation: BD stands for blood draw.

We used sequential readouts of the Roche Elecsys Anti-SARS-CoV-2 nucleocapsid assay (*11*) before (blood draws 8, 9) and at the end of (blood draw 10) the Omicron BA.1/2 wave to infer the cumulative rate of infections and re-infections during the Omicron BA.1/2 wave among the cohort populations. The inference of primary and repeat infections with Omicron was based on the dynamics of sequential serologic assay readouts, after calibrating the method on the Delta epidemic wave, for which we had both serology and PCR data. The analysis was performed on a subset of 905 of 1,200 cohort participants with complete serum specimens collected during the relevant Delta and Omicron periods (details of the serologic inference and calibration are described in Material and Methods Section 3). We ascertained infections predating the Omicron BA.1/2 wave based on both variant-specific PCR tests and serology, as detailed in a prior study ((*10*), covering the period through blood draw 8, or mid-September, 2021). All COVID-19 vaccinations were recorded, although the vaccine coverage remained low in this population, with <20% individuals receiving 1 or more doses prior to the emergence of the Omicron variant at both sites. In Table 1, we summarize the infection attack rate during the Omicron BA.1/2 epidemic wave by age, sex, rural and urban site, underlying medical conditions, and prior SARS-CoV-2 exposure.

**Table 1.**
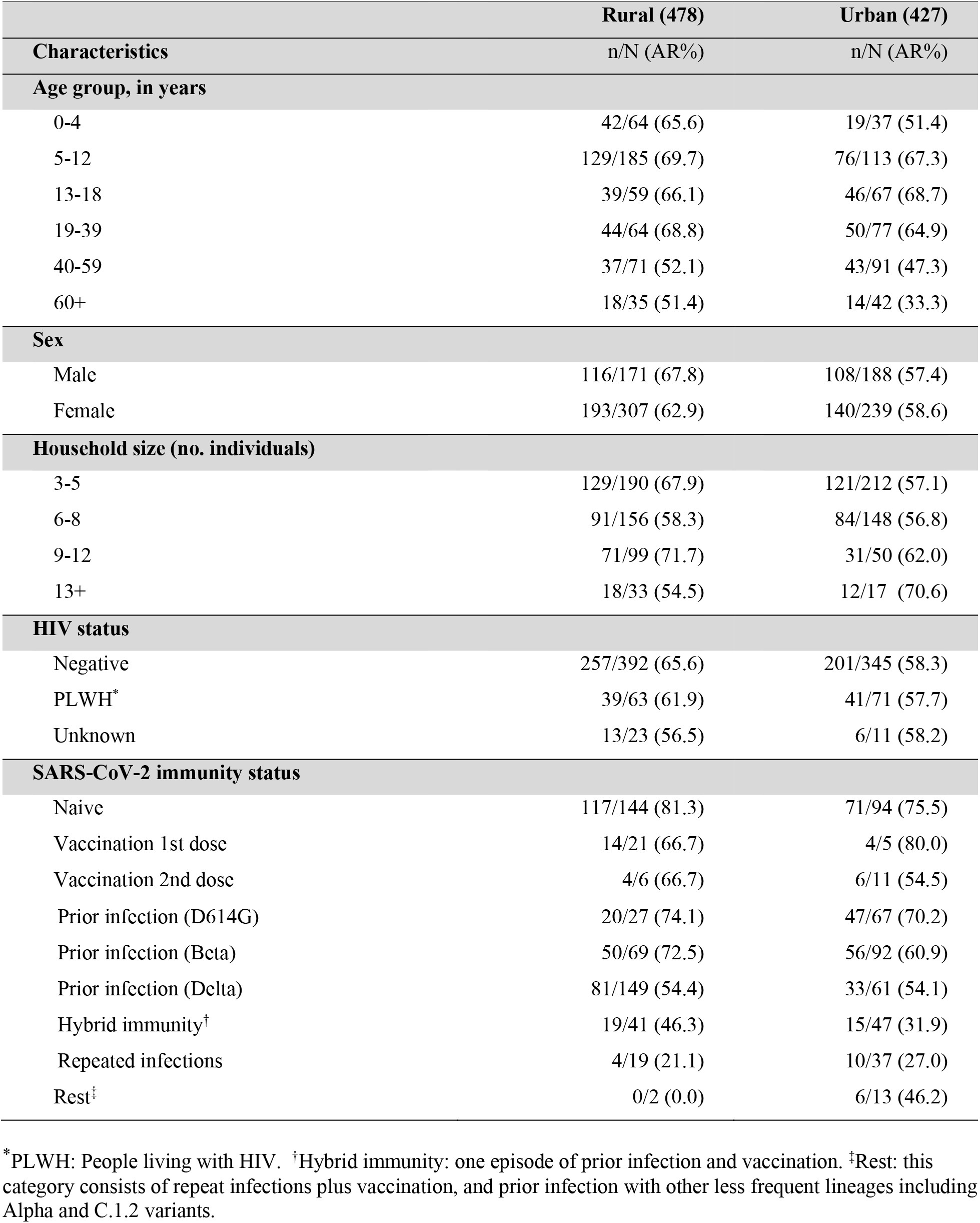
Omicron BA.1/2 infection attack rate (AR) in rural and urban cohorts of South Africa based on 905 participants with complete serologic information (75% of the total 1200 participants).

|  | **Rural (478)** | **Urban (427)** |
| --- | --- | --- |
| **Characteristics** | **n/N (AR%)** | **n/N (AR%)** |
| **Age group, in years** |  |  |
| 0-4 | 42/64 (65.6) | 19/37 (51.4) |
| 5-12 | 129/185 (69.7) | 76/113 (67.3) |
| 13-18 | 39/59 (66.1) | 46/67 (68.7) |
| 19-39 | 44/64 (68.8) | 50/77 (64.9) |
| 40-59 | 37/71 (52.1) | 43/91 (47.3) |
| 60+ | 18/35 (51.4) | 14/42 (33.3) |
| **Sex** |  |  |
| Male | 116/171 (67.8) | 108/188 (57.4) |
| Female | 193/307 (62.9) | 140/239 (58.6) |
| **Household size (no. individuals)** |  |  |
| 3-5 | 129/190 (67.9) | 121/212 (57.1) |
| 6-8 | 91/156 (58.3) | 84/148 (56.8) |
| 9-12 | 71/99 (71.7) | 31/50 (62.0) |
| 13+ | 18/33 (54.5) | 12/17 (70.6) |
| **HIV status** |  |  |
| Negative | 257/392 (65.6) | 201/345 (58.3) |
| PLWH* | 39/63 (61.9) | 41/71 (57.7) |
| Unknown | 13/23 (56.5) | 6/11 (58.2) |
| **SARS-CoV-2 immunity status** |  |  |
| Naive | 117/144 (81.3) | 71/94 (75.5) |
| Vaccination 1st dose | 14/21 (66.7) | 4/5 (80.0) |
| Vaccination 2nd dose | 4/6 (66.7) | 6/11 (54.5) |
| Prior infection (D614G) | 20/27 (74.1) | 47/67 (70.2) |
| Prior infection (Beta) | 50/69 (72.5) | 56/92 (60.9) |
| Prior infection (Delta) | 81/149 (54.4) | 33/61 (54.1) |
| Hybrid immunity† | 19/41 (46.3) | 15/47 (31.9) |
| Repeated infections | 4/19 (21.1) | 10/37 (27.0) |
| Rest‡ | 0/2 (0.0) | 6/13 (46.2) |

\*PLWH: People living with HIV. †Hybrid immunity: one episode of prior infection and vaccination. ‡Rest: this category consists of repeat infections plus vaccination, and prior infection with other less frequent lineages including Alpha and C.1.2 variants.

### Increasing population immunity and changing immunologic landscape over four epidemic waves among the PHIRST-C cohort population

Infection attack rates of the Omicron BA.1/2 subvariants were substantially higher than those of previously circulating variants in both study sites. In the rural site, infection rates for pre-Omicron variants were 8.2% (95% CI 5.7% – 11.0%) for D614G, 20.7% (95% CI 17.1% – 24.3%) for Beta and 38.7% (95% CI 34.3% – 43.1%) for Delta, with higher attack rates in each successive wave (Figure 2A, left panel). In the urban site, infection attack rates were more similar among pre-Omicron variants: 25.8% (95% CI 21.6% – 30.0%) for D614G, 32.1% (95% CI 27.7% – 36.5%) for Beta, 23.2% (95% CI 19.2% – 27.2%) for Delta (Figure 2B, left panel). Differences in infection attack rates between the urban and rural sites reflect the spatial heterogeneity of SARS-CoV-2 circulation in South Africa. Attack rates for the Omicron BA.1/2 subvariants reached 64.6% (95% CI 60.4% – 68.9%) in the rural site (Figure 2A, left panel) and 58.1% (95% CI 53.4% – 62.8%) in the urban site (Figure 2B, left panel).

**Fig. 2.**
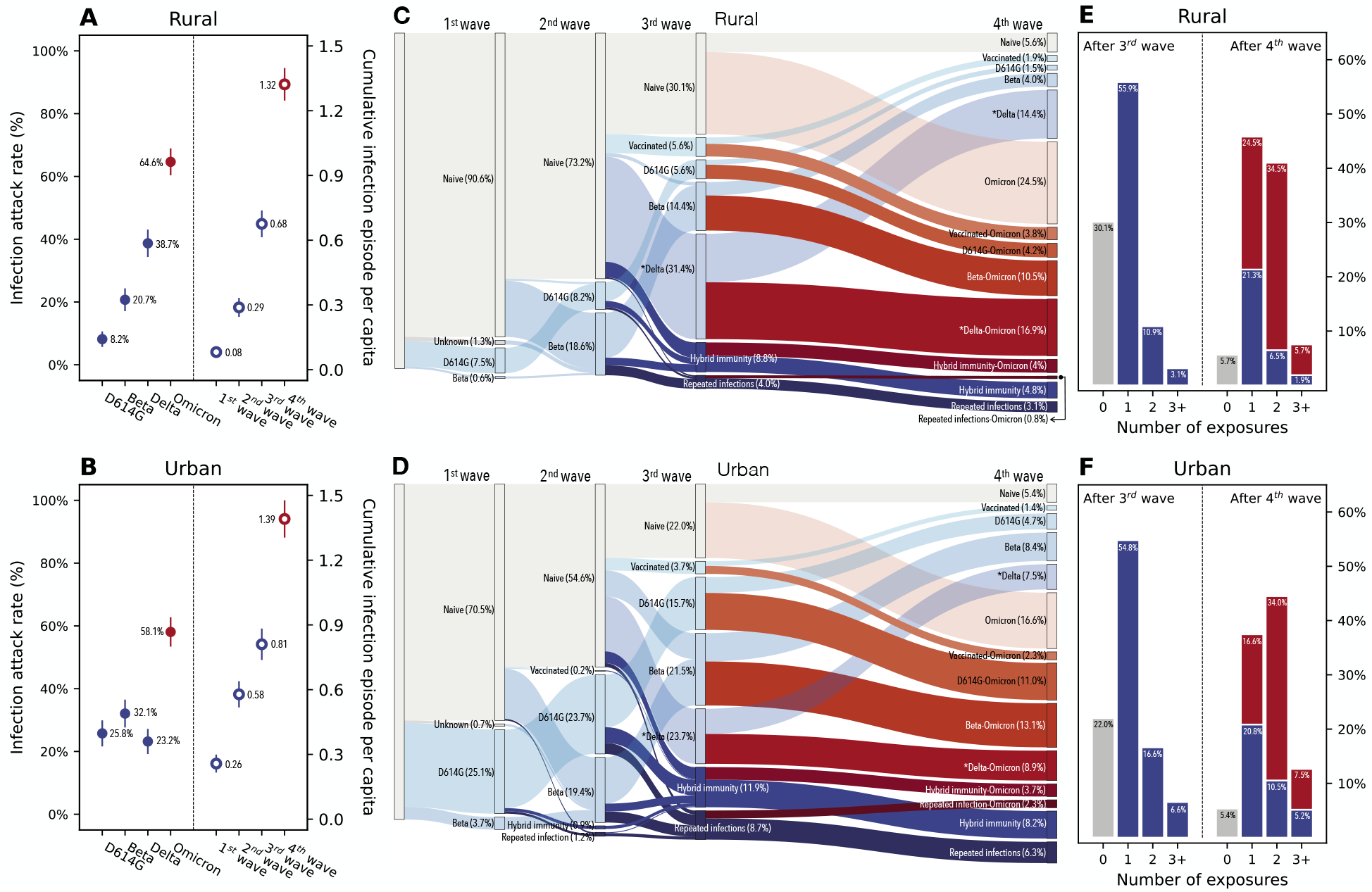
SARS-CoV-2 infection attack rates and shifts in immunologic landscape. (A) Infection attack rates in the rural site by variant type (left) and the cumulative number of infection episodes per capita after each epidemic wave (right). The end of the 1^st^ wave is marked by blood draw 2, 2^nd^ wave is marked by blood draw 5, 3^rd^ is marked by blood draw 8, 4^th^ is marked by blood draw 10. (B) Same as (A) but for the urban site. (C) Sankey diagram demonstrating the distribution of different type of immunologic exposures (including vaccination and infection) in the population of the rural site after each epidemic wave and the transition of immunologic exposures in-between waves. Gray color represents SARS-CoV-2 immunologic naïve individuals; blue shades represent non-Omicron exposures; red shades represent Omicron exposures; darker colors represent repeat exposures while transparent shading represents primary exposures. (D) same as (C) but for the urban site. (E) The distribution of the population by the number of SARS-CoV-2 exposures experienced after the 3^rd^ epidemic wave (left) or after the 4^th^ epidemic wave (right). Each infection/vaccine dose is counted as 1 exposure. Gray bars represent naïve individuals; blue bars represent individuals who have been infected by pre-Omicron variants or received SARS-CoV-2 vaccinations; red bars represent individuals who have been infected by Omicron. ^*^In additional to Delta, here also includes other less frequent lineages including other lineages including Alpha and C.1.2 variants.

Prior to the emergence of Omicron, in September 2021, the cumulative infection rate (including reinfections), was 67.6% (95% CI 61.4% – 73.8%) in the rural site and 81.0% (95% CI 73.8% – 88.3%) in the urban site (Figure 2 A and B, right panels), with primary infections accounting for the majority of all infections in both sites (Figure 2 E and F, left panels). In contrast, reinfections were predominant with Omicron BA.1/2, with primary infections representing only 37.9% (95% CI 32.5% – 43.3%) and 28.6% (95% CI 23.0% – 34.3%) of all Omicron BA.1/2 infections in the rural and urban site, respectively (Figure 2 E and F, right panels). As a result, cohort participants had experienced an average of 1.32 (95%CI 1.25 – 1.40) and 1.39 (95% CI 1.31 – 1.48) SARS-CoV-2 infection episodes in the rural and the urban site, respectively, approximately 2 years into the pandemic (Figure 2A-B).

Longitudinal rRT-PCR and serologic follow up at the individual level, combined with household-level random sampling scheme, provide a unique opportunity to track the history of SARS-CoV-2 immunizing events in the cohort populations, including vaccination (partially or fully) and infection. Figure 2 C and D are Sankey diagrams tracking the transition of different SARS-CoV-2 exposure histories after each of the four epidemic waves. The first three epidemic waves were dominated by primary exposures to D614G, Beta, and Delta variants. In addition, less than 20% of the population was vaccinated (most during the Delta wave) by either the Janssen Ad26.COV2.S or the Pfizer–BioNTech BNT162b2 vaccines prior to the emergence of Omicron. At the end of the third wave, only 30.1% and 22.0% of the population remained naïve to SARS-CoV-2 in the rural and the urban site, respectively. Most of the population had experienced a single SARS-CoV-2 exposure prior to Omicron’s emergence (55.9% in the rural and 54.8% in the urban site), while a minority had two or more exposures (14.0% and 23.2% in the rural and urban site respectively, Figure 2 E-F). This observation is in line with the finding of a durable immune protection conferred by prior infection and vaccination in the pre-Omicron era (*10*). In contrast, the large contribution of re-infections during the Omicron wave shifted the population immune landscape towards a dominance of repeat exposures (52.1% of individuals in rural site and 60.4% in the urban site had experienced more than one exposure, whether prior infection or vaccination, after the fourth epidemic wave). The high ratio of reinfections to primary infections observed during the Omicron wave is in line with a high level of population immunity predating Omicron BA.1/2’s arrival, combined with this variant’s immune evasion properties (ratio of reinfection to infection of 1.41 in the rural site and 2.04 in the urban site, Figure 2 E-F). As a result, the Omicron BA.1/2 wave left behind a heterogeneous immunological landscape, with population subgroups characterized by distinct exposure histories, and no exposure category accounting for more than 25% of the population (Figure 2 C-D).

### Risk factors, increased infectivity and immune evasion of the Omicron BA.1/2 variant, relative to Delta

Next, we fitted a chain-binomial household transmission model to the inferred serologic infections to contrast the characteristics of the Delta and Omicron variants, including the role of age, sex, household size, and prior exposure history (see Material and Methods Section 4 for details). We further differentiated the risk of transmission of primary infections and vaccine breakthroughs or reinfections, as well as the risk of acquiring infection within the household and from the community (stratified by age, sex and cohort site). We found that the Omicron BA.1/2 variant was more than twice as likely to transmit as the Delta variant (odds ratio Omicron vs. Delta: 2.35, 95% CI 2.11 – 2.62), after controlling for other risk factors (Figure 3). Vaccine breakthroughs and reinfections were 41% less likely to transmit than primary infections (odds ratio 0.59, 95% CI 0.47 – 0.74), suggesting a transmission reduction effect of prior immunity, in line with other findings (*12*).

**Fig. 3.**
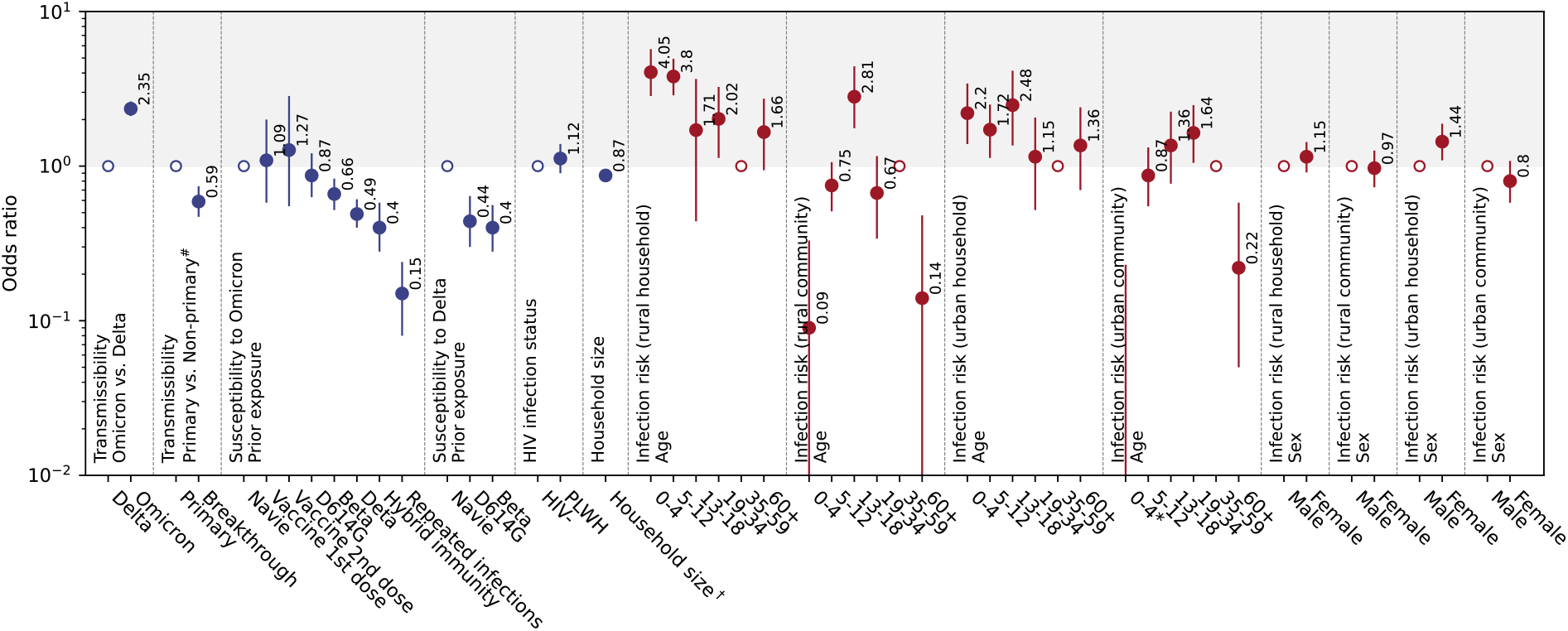
Risk factors associated with SARS-CoV-2 Omicron BA.1/2 and Delta infection. Odds ratios (adjusted after controlling for other risk factors, see Material and Methods Section 4 for details) were estimated by a chain binomial model where the infection status was inferred from serology. Abbreviation: PLWH: persons living with HIV. Category “Unknown” for “HIV infection status” and category “Rest” in “Prior exposure” (Table 1) were included in the model but omitted here due to small sample size in the strata. ^*^0-4 age group have odds ratio point estimate less than 0.01, thus not shown in the figure. ^#^Non-primary infections represent repeat/breakthrough infections. ^†^Household size denotes the number of household members within a household and is analyzed as a continuous variable.

Prior work has demonstrated Omicron BA. 1/2’s significant immune escape properties (*13–17*). In our data (Figure 3), the most recent prior infection with the Delta variant still conferred significant residual protection against Omicron (odds ratio 0.49, 95% CI 0.40 – 0.61), but protection declined from earlier infections: the odds ratio of prior Beta infection (2^nd^ wave) was 0.66 (95% CI 0.52 – 0.83), while a prior D614G infection (1^st^ wave) did not offer significant protection (0.87 (95% CI 0.63 – 1.21)). In contrast, prior Beta and D614G infections conferred a higher level of protection against Delta (odds ratios: 0.40 (0.28 – 0.56) and 0.44 (0.30 – 0.64), respectively) than against Omicron. Neither one nor two doses of vaccine showed significant protection against Omicron (Figure 3), despite >90% being administered within 5 months of Omicron’s emergence. It is also worth noting that <5% of participants (43/905, Table 1) had received one or two doses of vaccine before Omicron, resulting in wide confidence intervals on the effects of vaccination, and suggesting limited statistical power in these strata. However, hybrid immunity, defined as a combination of one prior infection (with any variant) with one or two doses of vaccines, conferred significant protection (odds ratio 0.40, 95% CI 0.28 – 0.58). Of note, repeat infections provided the strongest protection against the Omicron BA.1/2 variant, reducing the risk of infection by 85% (odds ratio 0.15, 95% CI 0.08 – 0.24).

In terms of behavioral and demographic factors, we found that the risk of transmission to household contacts (per-contact risk) decreased significantly with household size (number of household members) (odds ratio 0.87, 95% CI 0.83 – 0.90, Figure 3). Interestingly, we found that the risk of acquiring infection within the household or from the community varied substantially by age, sex and study site. Pre-school children aged 0-4 years had much higher risk of acquiring infection within the household relative to adults 35-49 years (odds ratio, urban site: 2.20, 95%CI 1.39 – 3.42; odds ratio, rural site: 4.05, 95% CI 2.84 – 5.71) but much lower risk of acquiring infection from the community (odds ratio, urban site: <0.001, 95% CI 0.00 – 0.23; odds ratio, rural site: 0.09, 95% CI 0.01 – 0.33, Figure 3). Similarly, individuals 60 years and older had very low risk of acquiring infection from the community (odds ratio, urban site: 0.22, 95% CI 0.05 – 0.58; odds ratio, rural site: 0.14, 95% CI 0.01 – 0.48). In the urban cohort, age group 19 – 34 years had the highest risk of acquiring infection from the community as compared to acquiring infection from the household (odds ratio: 1.64 with 95% CI 1.05 – 2.48), and so did age group 13 – 18 years in the rural site (odds ratio: 2.81 with 95% CI 1.76 – 4.42). Females had a significantly higher risk of infection within the household relative to males in the urban site (odds ratio female vs. male: 1.44, 95% CI 1.09 – 1.88) but there was no difference in the rural site.

### Comparing the disease severity of Omicron with that of earlier variants

Estimating the severity of Omicron BA.1/2 remains difficult due to profound changes in case reporting and the impact of prior exposures on clinical presentation, relative to prior pandemic waves. We compared our infection attack rate estimates with surveillance data to estimate the extent of under-reporting (*18*). The cumulative SARS-CoV-2 incidence rate reported by the surveillance system for the Omicron wave was 0.54 per 100 individuals in the health district of the rural site (Ehlanzeni District) and 0.76 per 100 in the urban site district (Dr Kenneth Kaunda District). The infection ascertainment rate was estimated at 0.84% in the rural site district and 1.31% in the urban site district, considerably lower than the ascertainment rates of prior waves (3% – 10%, *10, 11*), indicating that the surveillance system captured only a very small fraction of all Omicron BA.1/2 infections.

In Figure 4A, we estimate the infection fatality ratio (IFR) of each epidemic wave in the urban site of the study, which is more representative of South Africa’s urbanized population. We used the in-hospital death rate reported to the COVID-19 National Hospital Surveillance (*19*) at the district level (as numerator, Figure 4B) and the age-specific infection rates estimated in the PHIRST-C urban cohort (as denominator, Figure 4C). We estimate that the IFR was 0.043% (95% CI 0.040% – 0.047%) during the Omicron BA.1/2 wave, significantly lower than in the prior three waves (0.15% (95% CI 0.13% - 0.17%) during the 1^st^ wave dominated by D614G, 0.36% (95% CI 0.30% – 0.46%) during the 2^nd^ wave dominated by Beta, 0.41% (95% CI 0.37% – 0.47%) during the 3^rd^ wave dominated by Delta).

**Figure 4:**
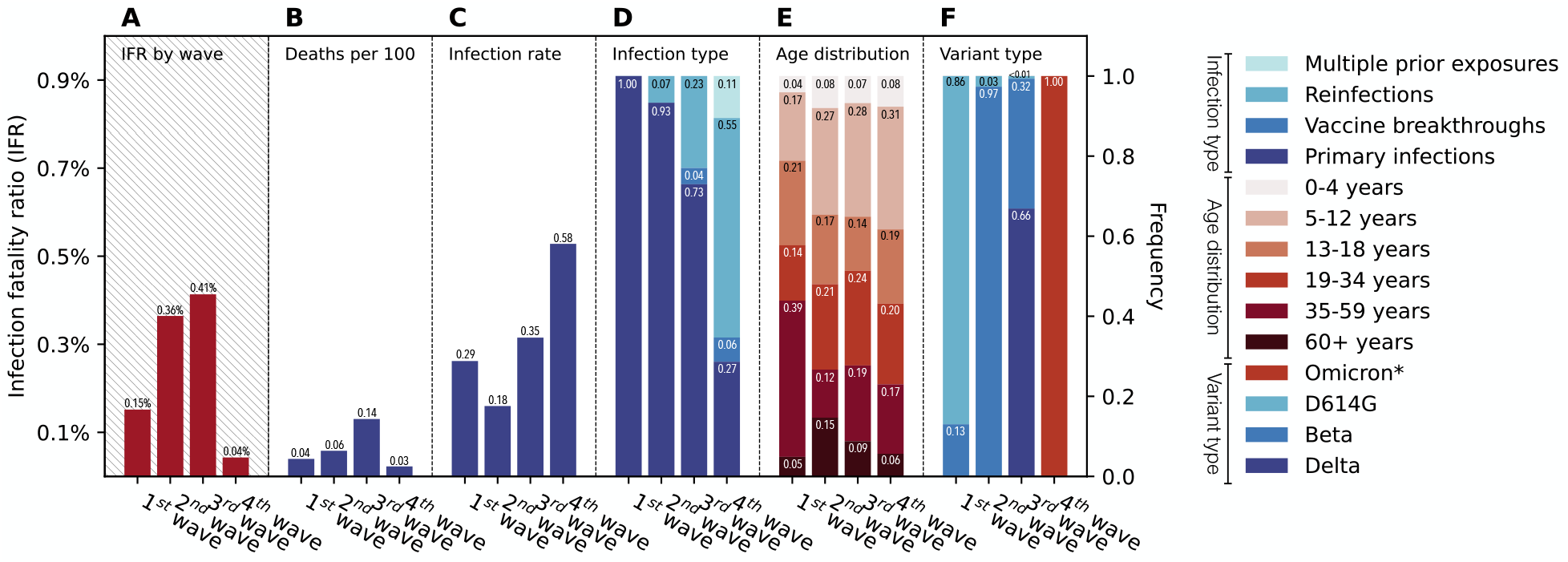
The infection fatality ratios and the factors associated with SARS-CoV-2 disease severity for different epidemic waves in the urban site’s district. (A) the estimated infection fatality ratio for each epidemic wave; (B) the mortality burden of each epidemic wave measured by the cumulative rate of in-hospital deaths per 100 individuals; (C) the infection attack rate of each epidemic wave in the North West based on the PHIRST-C urban cohort, assuming that the urban cohort population is representative of the population of the North West Province; (D) The wave-specific distribution of infection types based on prior exposure histories, including primary infection, vaccine breakthroughs (1 or 2 doses of vaccines), reinfections (infection after one prior infection), and multiple prior exposures (infection with two or more prior infections or a mixture of prior infection and vaccination). (E) The wave-specific age distribution of infections. (F) The wave-specific distribution of variant type among infections. Panel B-F share the same axis on the right. *For the 4^th^ wave, we could not confirm variant type by variant-specific rRT-PCR or sequencing, however, judging from the timing of emergence and dominance of Omicron in South Africa in late November 2021, we assumed here that all infections during the 4^th^ wave were due to Omicron BA.1/2 variants.

In Figure 4 D-F, we deconstruct the infection profile of each epidemic wave to highlight the role of key factors affecting severity. Prior immunity has an important role in shaping IFR, with the fraction of primary infections decreasing with each new epidemic wave, from 100% during the 1^st^ wave to 93%, and 73% and 29% during the Beta (2^nd^) and Delta (3^rd^) and Omicron BA.1/2 (4^th^) waves (Figure 4D). Notable changes in the age patterns of infections in different waves have also affected the IFR. In particular, the second wave had the highest proportion of infections in individuals 60 years and older (14.7%), followed by the 3^rd^ wave (8.8%), 4^th^ wave (6.9%), and 1^st^ wave (4.9%). Finally, each epidemic wave was dominated by a different variant (Figure 4D), and variations in the intrinsic severity of each variant would also impact the observed variations in IFRs across different epidemic waves.

## Discussion

Our findings indicate that the Omicron BA.1/2 wave had significantly higher attack rates than any of the previously circulating variants in South Africa, infecting 58-65% of the population in our urban and rural cohorts, despite high levels of pre-existing population immunity. Reinfections and vaccine breakthrough infections accounted for the majority of Omicron BA.1/2 infections, likely attributable to a minority of population remaining naïve prior to the Omicron BA.1/2 wave and to Omicron BA.1/2’s immune-escape properties (*16, 20, 21*). Additionally, a household transmission model estimated that participants were more than twice as likely to get infected during the Omicron BA.1/2 wave than the Delta wave, after adjustment for prior immunity and other factors. These findings support that the fitness advantage of Omicron BA.1/2 over Delta was not solely due to immune escape but also higher intrinsic transmissibility (*22*). However, relaxing nonpharmaceutical interventions on high infection attack rate during the Omicron wave likely also contributing to the higher infection risk during the Omicron BA.1/2 wave (vs. Delta wave), as prior to the 4^th^ epidemic wave (October 1 2021), the national COVID-19 measures were tuned down to Alert Level 1, the lowest level in South Africa’s COVID-19 alert system (*23*). It is worth noting that our serologic estimates of Omicron infection attack rates and the large contribution of reinfections and vaccine breakthroughs are in close agreement with earlier model projections for South Africa (in the urban site, the infection attack rate was estimated at 58.1% by serology vs. 44% - 81% by model projections, while the proportion of reinfections was 71% by serology vs. 49% - 72% by model projections (*10*)). These projections were based on a dynamic transmission model calibrated to the PHIRST-C urban site population and solely relied on epidemiological evidence of Omicron’s growth advantage and immune protection reduction (*10*). The concordance between model projections and post-projection serologic surveys highlights how mathematical modelling can synthetize routine surveillance data and detailed cohort information to anticipate epidemic size and other dynamical features of public health relevance, months before serologic surveys become available.

A unique feature of our cohort populations includes intense monitoring since the first SARS-CoV-2 epidemic wave, with nasal swab collection at a twice weekly frequency, combined with frequent serum specimen collection (*10*). Combining genomic sequencing, variant-specific rRT-PCRs, serologic testing, as well as information on vaccine administration, we had the unusual opportunity to track each individual participant’s SARS-CoV-2 antigen exposure history in chronological order. Random sampling of households ensured that the immune exposure distribution within these cohorts reflected the immunologic landscape of the broader population at the study sites. After the third epidemic wave dominated by Delta, and prior to Omicron’s emergence, more than 60% of the population at both sites had been infected by and/or vaccinated against SARS-CoV-2 at least once, with limited occurrence of reinfections (Figure 2). While there are different hypotheses addressing the evolutionary origins of Omicron BA.1’s emergence and this variant’s unusual number of mutations (*24*), the rising level of SARS-CoV-2 exposure in South Africa in 2020-2021 would be expected to promote the fitness advantage of immune escape variants over earlier variants. Globally, especially in high income countries, a similar shift in the immunologic landscape appears to have occurred, mainly via increased vaccine coverage, allowing Omicron to out-compete other variants and rapidly reach global dominance within 3 months of initial detection in Southern Africa (*5*). In addition to immune escape, our data also support a significant enhanced transmissibility advantage of Omicron BA.1/2, which would also contribute to rapid replacement of pre-existing variants.

After the Omicron BA.1/2 epidemic wave in South Africa, only 5.7% and 5.4% of the cohort population remained naïve to SARS-CoV-2 in the rural and urban sites respectively, and past immune histories in the rest of the population were highly diverse. Remarkably, the immunologic landscape of the population has become highly fragmented, with no single SARS-CoV-2 exposure category representing more than 25% of the population (Figure 2). For instance, among 379 individuals (across both study sites) who experienced a single SARS-CoV-2 exposure, 49.6% (188/379) were infected by the Omicron variant and the rest were exposed to pre-Omicron variants or vaccines. These two population groups primed by different antigens will likely have different antibody responses as the sera of individuals primed by pre-Omicron variants or vaccines poorly neutralize the Omicron BA.1/2 variant and vice versa (*15, 25*). Among individuals that were primed by different types of pre-Omicron variants or vaccines, differences in antibody responses are also expected, though they would be less prominent than the differences between individuals primed by the Omicron vs. pre-Omicron variants (*26*).

After the 4^th^ wave, among individuals who experienced two or more SARS-CoV-2 exposures, most had experienced Omicron BA.1/2 reinfections or vaccine breakthroughs with Omicron (80.3%, Figure 2 C-D). As SARS-CoV-2 moves into the endemic phase, multiple antigen exposures will become the norm, and these complex exposure histories will define population immune pressure to select new variants. We can see some of this play out already in the rapid succession of BA.1/2 and BA.4/5 waves, with in vitro experimental and computational predictive approaches informing which mutations are likely to become successful. Deep mutational scanning experiments of the SARS-CoV-2 receptor binding domain suggest that further mutations of Omicron BA.1/2 at sites 452 and 486 could lead to further immune evasion against antibodies isolated from individuals who had pre-Omicron and Omicron BA.1 breakthrough infections (*20, 27, 28*). Our study suggests that after the Omicron BA.1/2 wave, the population was dominated by Omicron BA.1/2 convalescent individuals (Omicron BA.1/2 breakthroughs/reinfections in particular), and hence the strongest immune pressure would arise from these individuals. Strikingly, mutations L452R and F486V have appeared in Omicron BA.4/5 lineages and these variants have now caused a fifth epidemic wave in South Africa. Interestingly, repeat infections by two or more pre-Omicron variants accounted for only a small fraction of the population after the third wave in our data, but these individuals retained great protection against Omicron BA.1/2.

As a large fraction of the population in South Africa (and globally) have experienced two or more antigenically distinct exposures involving Omicron lineages, prior variants, and/or vaccination episodes, the potential impact of immune imprinting needs to be closely monitored. The effects of imprinting have already been observed, where, for instance, individuals primed by spike-based vaccines have a more spike-focused immune response, compared to individuals primed by pre-Omicron infections (in which case, priming also includes non-spike proteins) (*29, 30*). It will be important to evaluate how imprinting by Omicron and pre-Omicron spike antigens differentially affects the immune response. Several studies have demonstrated that the antibody response after Omicron BA.1 breakthrough infection is dominated by recall memory B-cells against conserved epitopes shared between pre-Omicron strains and Omicron BA.1/2. Over reliance on immune memory may impact the breadth and strength of antibody and B-cell immunity as new antigenically drifted variants arise (*15, 20, 31, 32*). With each new variant wave, and vaccine reformulation anticipated for later in 2022, imprinting could play a role in shaping future disease trajectories. Ensuring the continuation of long-term cohort studies is particularly key to answering this question and guiding future vaccine updates.

Comparing our serology-based infection estimates with surveillance data indicates that only the “tip of the iceberg” of all SARS-CoV-2 infections (less than 1%) were captured by routine surveillance during the Omicron BA.1/2 epidemic wave in South Africa. On the other hand, weekly case surveillance accurately captured the speed of Omicron’s spread, and mathematical models calibrated to these data generated early projections of epidemic size that are comparable to serologic estimates (*10*). Despite a particularly high rate of infection with the Omicron BA.1/2 wave, there were fewer in-hospital deaths reported during this wave than in all prior waves in South Africa, suggesting a marked reduction in the overall infection fatality ratio (Figure 4). The lower estimated disease severity of Omicron BA.1/2 needs to be interpreted in the context of changes in infection demographics, circulating variant, and immunologic landscape across waves (*33, 34*). Firstly, the intrinsic severity of Omicron BA.1/2 could be lower than that of prior variants, especially when compared to Delta, as suggested by observational studies (*35, 36*). In addition, *in vitro* studies support Omicron’s limited ability to infect the lung (*37*), which could be a potential mechanism for Omicron BA.1/2’s observed attenuated severity. Secondly, the level of prior immunity was much higher during the Omicron BA.1/2 epidemic wave, as shown in our data. If prior immunity confers significant protection against SARS-CoV-2 deaths, as many studies show, then the high level of prior immunity at the start of the Omicron BA.1/2 wave would also contribute to the perceived attenuation of disease severity. It is also worth noting that “survival bias” could further underestimate the crude IFR, whereby frail individuals may have succumbed to earlier waves and left a healthier population at the start of the Omicron wave (*38–40*). Finally, the age profile of infections could also influence the overall IFR, as the risk of SARS-CoV-2 death increases exponentially with age, with most COVID-19 deaths concentrated in the population 60 years and older (*41*). The proportion of infections among individuals 60 years and older was 6.9% during the Omicron wave, intermediate between prior waves. Future cohort analyses of Omicron BA.1/2 disease severity need to carefully adjust for age, prior infection and vaccination, and potential observational biases.

Our study has several limitations. First, Omicron BA.1/2 infections were ascertained by serology without confirmation by rRT-PCR, and infections may have been misclassified due to imperfect sensitivity and specificity of the serologic approach. We trialed the approach during the Delta wave, by comparing serology with rRT-PCR results, and found appropriate sensitivity and specificity (Figure S4). Refinement of the serologic analysis and/or improved serologic assays more sensitive to waning (*42*) could further reduce potential misclassification. Second, the lack of virologic samples hindered differentiation between the Omicron BA.1 and BA.2 lineages, which may have different epidemiological properties in terms of transmissibility, immune evasion, and disease severity. Lastly, our study was concentrated on a relatively confined geographic scale and findings may not be representative of the broader population in South Africa.

In conclusion, our study suggests the massive scale of the Omicron BA.1/2 wave of infections in South Africa, orders of magnitude larger than the observations based on surveillance data. Reinfections and vaccine breakthrough infections dominated this wave due to combined effects of Omicron BA.1/2’s immune evasion properties and a high proportion of the population with pre-existing immunity. The overall disease severity of Omicron BA.1/2 was much lower than that of pre-Omicron waves, reflecting the contribution of attenuated intrinsic severity of this variant, but also changes in the age patterns of infections, and the impact of prior immunity. The remarkably diverse population immunologic landscape left by successive waves of SARS-CoV-2 will affect the epidemiological success of future variants. Our study sites are uniquely located in a region which has witnessed the emergence of key SARS-CoV-2 variants including Beta, Omicron BA.1, BA.4 and BA.5. As we reach the endemic phase of SARS-CoV-2, longitudinal studies will continue to be important to monitor shifts in population immunologic landscape and provide important context for understanding the fitness advantage and adaptive evolution of current and future variants.

## Data Availability

The investigators welcome enquiries about possible collaborations and requests for access to the data set. Data will be shared after approval of a proposal and with a signed data access agreement. Investigators interested in more details about this study, or in accessing these resources, should contact the principle investigator, Prof Cheryl Cohen, at NICD (cherylc{at}nicd.ac.za).

## Acknowledgment

We thank Dr. Michael P. Busch from Vitalant Research Institute and the University of California San Francisco for helpful feedback and suggestions. The findings and conclusions in this report are those of the authors and do not necessarily represent the official position of the NIH or the U.S. Centers for Disease Control and Prevention.

## Author contributions

KS, ST, JK, AvG, MLM, NW, JM, NAM, KK, LL, CV, CC designed the experiments. CC, JK, and ST accessed and verified the underlying data. ST, JK, AvG, MLM, NW, JNB, JM, MC, NAM, KK, LL, JdT, TM, CC collected the data and performed laboratory experiments. KS, ST, JK, AvG, MLM, NW, JNB, JM, MC, NAM, KK, LL, JdT, TM, CV, and CC analyzed the data and interpreted the results. KS, ST, JK, AvG, CV, and CC drafted the manuscript. All authors critically reviewed the Article. All authors had access to all the data reported in the study.

## Ethics statement

The PHIRST-C protocol was approved by the University of Witwatersrand Human Research Ethics Committee (Reference 150808) and the U.S. Centers for Disease Control and Prevention’s Institutional Review Board relied on the local review (#6840). The protocol was registered on clinicaltrials.gov on 6 August 2015 and updated on 30 December 2020 (https://clinicaltrials.gov/ct2/show/NCT02519803). Participants receive grocery store vouchers of ZAR50 (USD 3) per visit to compensate for time required for specimen collection and interview.

## Supplementary Materials

### Materials and Methods

#### 1. Cohort design and the timing of serum sample collection for the Omicron BA.1/2 wave

The study builds on prior work on the PHIRST-C cohorts (PHIRST-C is an acronym for **P**rospective **H**ousehold study of SARS-CoV-2, **I**nfluenza and **R**espiratory **S**yncytial virus community burden, **T**ransmission dynamics and viral interaction in South Africa, PHIRST-C, where “**C**” stands for COVID-19). Details on these South African cohorts have been previously described elsewhere (*9, 10*). To briefly summarize, the PHIRST-C cohorts consist of 114 households (638 participants) in a rural site located in Agincourt, a rural community in Mpumalanga Province, and 108 households (557 participants) in an urban site located in Jouberton township, Matlosana, North West Province. These cohorts were subject to an intense period of follow-up, from July 2020 to August 2021, during which nasal swab specimens were collected twice-weekly and a total of 7 blood draws (BDs) were collected at enrollment and then approximately every two months. We used these data to reconstruct the prior exposure history of each cohort individual before Omicron arose, and to calibrate a serology-based approach to infer infections during the Omicron period. After the intense follow-up period concluded, three additional BDs were collected at both study sites (Figure 1), with BD 8 collected in mid-September 2021 (the end of Delta wave), BD 9 in mid-November 2021 (at the time Omicron BA1/2’s emergence in Southern Africa, but prior to the rise in reported cases), and BD 10 in late March 2022 (at the end of South Africa’s Omicron BA.1/2 wave). We used these data to infer the Omicron infection status of each cohort participant.

#### 2. Laboratory methods

The laboratory methods of the intense follow-up period have been described in detail in prior study by Cohen et. al. (*9*). Serum specimens collected at blood draws 8, 9, and 10 follow the same protocol as those for prior blood draws detailed in (*9*): briefly, serum specimens were collected using venous blood, centrifuged into serum separator tubes, refrigerated immediately and transported to the NICD laboratories. According to manufacturer instructions, aliquots of prespecified volume were tested for the presence of SARS-CoV-2 antibodies by the Roche Elecsys Anti-SARS-CoV-2 nucleocapsid (N) assay (*43*).

#### 3. Inference of SARS-CoV-2 Omicron infections among PHIRS-C participants based on the antibody trajectory estimated from pre- and post-Omicron BA.1/2 (4^th^) wave blood draws

##### 3.1 Overview

As the PHIRST-C intense follow-up period ended in August 2021, individuals were no longer tested by rRT-PCR at twice-weekly frequency, thus Omicron BA.1/2 infections in the cohort population can only be ascertained through serology. We borrowed a paired sera approach from prior influenza studies, where serum samples are collected before and after the epidemic wave, with the rise in antibody titers between the pre- and post-wave sera used as a marker of influenza infection. This approach has been the gold standard to ascertain influenza infection attack rate, as prior exposures were common in the population (*44*). To identify SARS-CoV-2 primary infections and reinfections during the Omicron BA.1/2 wave at both sites, we relied on the serial serologic results of BD 8, 9, (pre-Omicron BA.1/2 wave) and BD 10 (post-Omicron BA.1/2 wave) by the Roche Elecsys Anti-SARS-CoV-2 nucleocapsid (N) assay (refer to as Roche anti-N here after). To capture both primary infections and reinfections occurring in-between BD 8, 9, and 10, rather than relying on a boost of pre-post season paired sera as the sole marker of infection (*44*), we finely categorize the serial BDs patterns based on the sequential seroconversion and boosting/waning patterns of the Roche anti-N assay readouts. To trial the approach, we relied on a study period surrounding the Delta wave where we had both periodic serology and twice-weekly Rt-PCR testing on all individuals. We matched BDs 8, 9, and 10 (4^th^ wave dominated by Omicron) with BDs 5, 6 and 8 (3^rd^ wave dominated by Delta) based on the similar timing of serum specimen collection with respect to the epidemic curves of these waves (Figure 1). We identified the serial serologic patterns most strongly associated with rRT-PCR confirmed SARS-CoV-2 infections during the Delta wave. We then generalized the serial serologic patterns to BDs 8, 9, and 10 to infer Omicron BA.1/2 infections during the 4^th^ epidemic wave.

##### 3.2 Categorization of serial serologic patterns during the 3^rd^ and 4^th^ epidemic waves in South Africa

The Roche anti-N is a commercial assay that was calibrated to detect recent and prior SARS-CoV-2 infections, based on the level of serum antibody against the SARS-CoV-2 N protein. The assay cutoff index (COI) above or equal to 1 marks seropositivity, while a COI below 1 is deemed seronegative (*43*). We assessed each participant’s serial serologic trajectory from pre- and post-wave serum specimens, measured by Roche anti-N COIs,. We used the assay seroconversion (anti-N COIs going from below 1 to above 1) as evidence of primary infection. We also used further rises in COI from a seropositive baseline (COI above 1) as a marker of reinfection, as a new exposure would be expected to generate anamnestic boosting of anti-N antibody levels above prior levels. We inferred SARS-CoV-2 primary infections and reinfections during the 3^rd^ epidemic wave based on participants’ serial serologic trajectory from pre- and post-wave serum specimen (measured in Roche anti-N COIs). We selected two BDs prior to the 3^rd^ (Delta) epidemic wave (BD 5 and 6) and one BD at the end of the 3^rd^ epidemic wave (BD 8) (Figure 1). We did not consider BD 7, because this offered an extra sampling time point that we would not have for the Omicron wave. We aimed to infer the occurrence of primary/repeat infections that occurred between May 1, 2021 (roughly the mid-point between BD 5 and BD 6) and BD 8, covering the majority of the 3^rd^ epidemic wave. It’s worth noting that we chose the mid-point between BD 5 and BD 6 as the start date of 3^rd^ wave inference to mirror the timing of the Omicron emergence in South Africa during the 4^th^ wave, which roughly falls in-between BD 8 and 9, in early November 2021.

As the majority of the 3^rd^ wave infections occurred in-between BD 6 and BD 8 (Figure 1), changes in the COIs from BD 6 to BD 8 are likely most informative of SARS-CoV-2 infections between the two blood draws. We categorized the sequential serologic patterns as follows, based on the Roche anti-N assay:

1. Seroconversion from BD 6 (seronegative, COI<1) to BD 8 (seropositive, COI≥1). This was likely due to a SARS-CoV-2 primary infection occurring between BD 6 and BD 8. However, sero-negativity at BD 6 could also correspond to a prior infection that had sero-reverted by BD 6. In this case, seroconversion from BD 6 to BD 8 would suggest a reinfection between BD 6 and BD8. If sero-reversion had occurred by BD6, then the individual should be seropositive at the earlier blood draw (BD5).
2. If a participant was already seropositive at BD 6, we hypothesized that a reinfection occurring between BD 6 and BD 8 would induce anamnestic “boosting” of the anti-N antibody level which could lead to an increase in BD 8’s COI when compared to BD 6’s. Evidence of reinfection is particularly strong if the COI of BD 6 was lower than the COI of BD 5, establishing a waning baseline prior to further exposure. In this case, serial COI is expected to follow a “V-shape” trajectory from BD5-8, as suggested by a prior study also using serial blood samples to detect reinfections (*45*). Establishing a waning baseline is especially informative for the Roche anti-N assay. A prior study suggests that a small fraction of individuals can show a gradual increase in Roche anti-N COI up to 4 months after SARS-CoV-2 infection (*46*), likely due to this assay using a dual-antigen antibody detection method tuned for high avidity antibodies (*47*). Thus “double boosting” at BD 5, BD 6 then BD 8 may not necessarily suggest reinfection, and a boosting threshold may be needed to differentiate strong boosting (most likely due to reinfection) from weak boosting (due to the nature of the Roche-N longitudinal kinetics or measurement error). We will formally test these serial serologic patterns on their predictability of SARS-CoV-2 infections through comparing them with rRT-PCR confirmed infections during the Delta wave (detailed below then in Material and Methods Section 3.3).

Building on the above logic, we start by categorizing the serial Roche anti-N COIs of BD 5, 6, and 8 into eight crude categories A, B, C, D, E, F, G, and H based on seroconversion and boosting/waning of the serial BDs’ COIs, specifically:

- Category A: Seronegative at each BD 5, 6, and 8. This pattern is consistent with no infection before and during Delta wave.
- Category B: Seropositive at BD 5 followed by decreasing COIs (waning) from BD 5 to BD 6 and further decline from BD 6 to BD 8. This suggests a prior infection before the Delta wave, and waning during Delta wave.
- Category C: Seropositive at BD 5 followed by a rise in COI from BD 5 to BD 6 (boosting) then a decline in COI from BD 6 to BD 8. This could be evidence of re-infection between BD 5 and BD 6; we further refine this logic to distinguish reinfection “boosting” from long-term COI increase due to infection prior to BD 5 or measurement noise (see below).
- Category D: Seronegative at BD 5 followed by seroconversion at BD 6 then declining COI from BD 6 to BD 8. This is evidence of a Delta infection between BD 5 and BD 6.
- Category E: Seropositive at BD 5 followed by rise in COI from BD 5 to BD 6 then further rise of COI from BD 6 to BD 8. This could be evidence of reinfection between BD 5 and BD 6 or reinfection between BD 6 and BD 8; we further refine this logic to distinguish reinfection “boosting” from long-term COI increase due to infection prior to BD 5 or measurement noise (see below).
- Category F: Seronegative at BD 5 followed by seroconversion at BD 6 and then boosting COI from BD 6 to BD 8. This could be evidence of primary infection between BD 5 and BD 6 and/or reinfection between BD 6 and BD 8; we further refine this logic to distinguish reinfection “boosting” from long-term COI increase due to infection prior to BD 6 or measurement noise (see below).
- Category G: Seropositive at BD 5 followed by waning COI from BD 5 to BD 6 and boosting COI from BD 6 to BD 8. This could be evidence of reinfection between BD 5 and BD 8; we further refine this logic to distinguish reinfection “boosting” from measurement noise (see below).
- Category H: Seronegative at BD 5 and BD 6 followed by seroconversion in-between BD6 and BD 8. This is evidence of primary infection between BD 6 and BD 8.

Figure S1 A-H visualize the serologic trajectory of BD5, 6, and 8’s Roche anti-N COIs for individuals in Categories A through H, respectively. The quantitative criteria for the crude serial serologic pattern categorizations are listed in Table S1 under “Categorization (crude)”.

A further rise of Roche anti-N COIs among seropositive individuals could be a marker for SARS-CoV-2 reinfections. To identify the degree of boosting most concordant with reinfection, we refine the crude categories listed above into finer categories by introducing a hyperparameter of boosting threshold *γ*>1: in a seropositive individual, a further increase of COI above the threshold *γ* may indicate a stronger signal of anamnestic boosting in antibody level due to reinfection, while boosting below *γ* is consistent with the slow long-term rise of antibody level from prior infection measured by the Roche anti-N assay (*46, 47*) and/or measurement noise (see next section for calibrating *γ* for the optimized sensitivity/specificity). This refined characterization of serial serologic patterns is applied to crude categories C, E, F, G (categories with further boosting from seropositive baseline), specifically:

- Category C is further divided into two sub-categories C_0_ and C_1_, where C_1_ requires BD_6_/BD_5_ > *γ* and C_0_ requires BD_6_/BD_5_ > *γ*.
- Category E is further divided into three sub-categories E_0_, E_1_, and E_2_, where E_2_ requires BD_8_/BD_6_ > *γ*, E_1_ requires BD_8_/BD_6_ > *γ* & BD_6_/BD_5_ > *γ*, and E_0_ requires BD_8_/BD_6_ > *γ* & BD_6_/BD_5_ > *γ*.
- Category F is further divided into to two sub-categories F_0_ and F_1_, where F_1_ requires BD_8_/BD_6_ > *γ* and F_0_ requires BD_8_/BD_6_ > *γ*.
- Category G is further divided into two sub-categories G_0_ and G_1_, where G_1_ requires BD_8_/BD_6_ > *γ* and G_0_ requires BD_8_/BD_6_ > *γ*.

The quantitative criteria for the refined subcategorization described above are also listed in Table S1 under the “Categorization (refined)” of the 3^rd^ epidemic wave. Similarly, we categorized serologic patterns in BDs 8, 9, 10 (with BD 8 matching BD 5, BD 9 matching BD 6, and BD 8 matching BD 10) based on quantitative criteria listed in Table S2. Figure S2 A-H visualize, from Category A through H respectively, the serologic trajectory of BD8, 9, and 10’s Roche anti-N COIs.

##### 3.3 Calibrating *γ* and selecting serial serologic patterns associated with SARS-CoV-2 primary infections and reinfections among PHIRST-C cohorts during the 3^rd^ epidemic wave in South Africa (intense follow-up period)

We optimized the *γ* value as well as serial serologic patterns that best differentiated between individuals with SARS-CoV-2 primary infections and reinfections from non-infected individuals. This analysis was based on infections occurring between the mid-point of BDs 5 and 6 and BD 8, corresponding to the 3^rd^ wave dominated by Delta, in the period where rRT-PCR was available. We scanned through *γ* values from 1 to 3 using a step size of 0.1. Under each *γ* value, we iterated through each of the 13 refined serial serology subcategories (A, B, C_0_, C_1_, D, E_0_, E_1_, E_2_, F_0_, F_1_, G_0_, G_1_, H, detailed in Table S1). We calculated the net contribution to the Youden’s J statistics if the pattern of interest was considered as a marker of infection (primary or reinfections) during the time window of interest (Youden’s J = sensitivity + specificity – 1, the higher the value of Youden’s J, the better the performance of the classification balancing both sensitivity and specificity). We then selected all serial serologic patterns with net-positive contribution to the Youden’s J statistics as markers for infection, while the rest of the 13 serial serologic patterns were markers of non-infection. This gave the best Youden’s J statistics under a given *γ* value. We found that at *γ*=1.4, we arrived at the highest Youden’s J statistics of 0.820 and good sensitivity (0.804) for reinfections (Figure S3) with patterns E_2_, F_1_, G_1_, H indicating SARS-CoV-2 infections while the rest (A, B, C_0_, C_1_, D, E_0_, E_1_, F_0_, G_0_) indicating absence of infections (Figure S4 & Table S1). We generalize this categorization of infections (E_2_, F_1_, G_1_, H) vs non-infections (A, B, C_0_, C_1_, D, E_0_, E_1_, F_0_, G_0_) to BDs 8, 9, 10 to infer infections during the 4^th^ (Omicron BA.1/2) wave.

#### 4. Chain-binomial SARS-CoV-2 household transmission model

Here we studied predictors and risk factors of SARS-CoV-2 infection during the Delta and the Omicron waves, in particular the contribution of prior immunity, demographic and medical characteristics, other infections in the household, and the risk of infection from the community. We used a chain-binomial household transmission model for SARS-CoV-2, as an extension of prior household models developed to study influenza transmission (*48, 49*). We jointly fitted the model to both the 3^rd^ (Delta dominated) and 4^th^ waves (Omicron BA.1/2), using serology-inferred infections as the outcome, as described in Section 3. Our model considers both community-acquired infections as well as multigenerational transmission within the household.

We denote 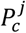 as the risk of individual *j* acquiring infection from outside the household (community); to model the risk of transmission between household contacts, we denote 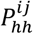 as the risk of an infected household contact *i* infecting household contact *j* in household *h*. We can express 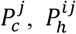 as:

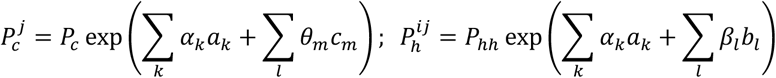

where:

- *P*_*c*_ denotes the baseline SARS-CoV-2 community infection risk.
- *P*_*hh*_ denotes the baseline risk of SARS-CoV-2 transmission between an infected and another uninfected household contact.
- *a*_*k*_ denotes risk factors *k* shared by community-acquired infection and household transmission, including, variant-specific transmissibility, variant-specific susceptibility due to prior exposure history, and HIV infection status (see Figure 3).
- *b*_*l*_ denotes household-specific risk factor *l* that could potentially influence household transmission, including transmissibility of primary vs breakthrough infections, household size, site-specific age & sex (see Figure 3).
- *c*_*m*_ denotes community-specific risk factor *m* that could potentially influence the acquisition of infection from the community, including site-specific age & sex (see Figure 3).

Let *h* denote a household, *i* an individual, and 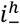 an individual *i* who remained infection-free during the Delta/Omicron waves in household *h*, while 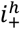 is an individual *i* who was infected in household *h* (as determined by serology). We can then express the probability of household member *i* escaping infection from all infected household contacts as:

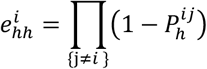

where {*j* ≠ *i*} represents all infected household contacts. We can also express the probability of household member i escaping infections from the community as:

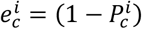

Thus, within household *h*, the likelihood of a household contact *i* being non-infected is given by:

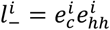

And the likelihood of an individual *i* being infected is given by:

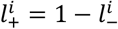

For household *h*, the loglikelihood of observing the infection status of all household contacts is given by:

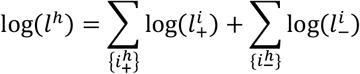

The overall likelihood of the observations across all households is given by:

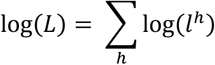

We used maximum likelihood method to optimize the function of log(*L*), and obtain point estimates of {*α*_*k*_}, {*β*_*l*_}, {*θ*_*m*_} and their corresponding confidence interval using likelihood ratio test.

**Table S1:**
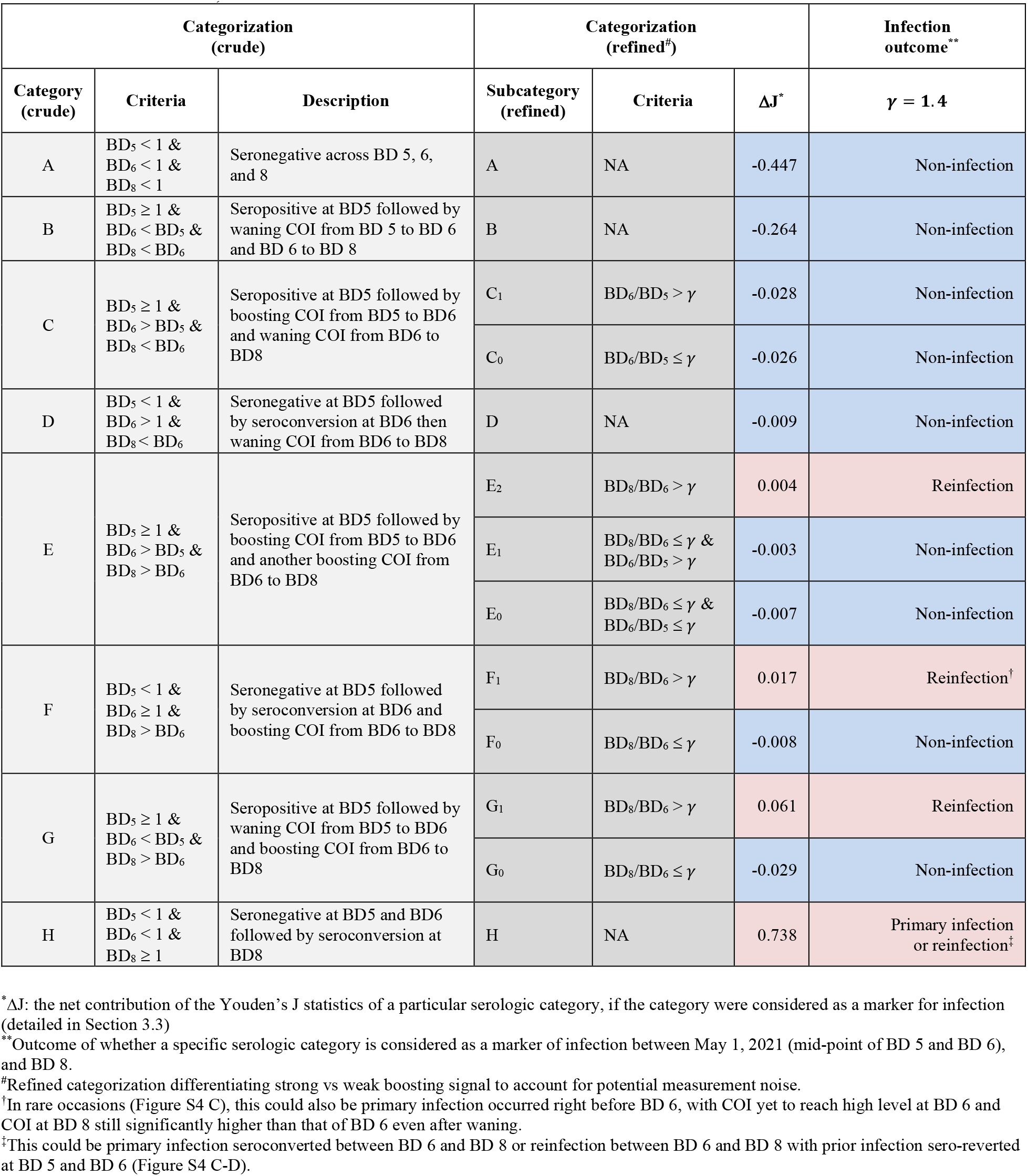
Categorization and infection outcome calibration of the 3^rd^ wave serial serologic patterns (BDs 5, 6, 8, Delta wave). The outcome is a SARS-CoV-2 infection occurring between the mid-points of BD 5 and BD 6, and BD 8. These categories are used to calibrate a serology-based approach to infer infections during a the period of intense cohort follow up period, when rRT-PCR confirmed infections are available (detailed in Material and Methods Section 3.2-3.3).

**Table S2:** Categorization of the 4^th^ wave serial serologic patterns (BDs 8, 9, 10, Omicron wave). The determination of infections outcomes (last column) for each subcategory of sequential serologic patterns A-H are based on the calibration detailed in Table S1. This approach captures infections and re-infections that occurred between the mid-point of BD 8 and BD 9, and BD 10, during the Omicron BA1/2 wave.

| Wave | Categorization (crude) |  |  | Categorization (refined<sup>#</sup>) |  | Infection outcome<sup>*</sup> |
| --- | --- | --- | --- | --- | --- | --- |
| 4<sup>th</sup> epidemic wave in South Africa | Category (crude) | Criteria | Description | Subcategory (refined) | Criteria | $\gamma = 1.4$ |
| | A | $BD_8 < 1$ &<br>$BD_9 < 1$ &<br>$BD_{10} < 1$ | Seronegative across BD 8, 9, and 10 | A | NA | Non-infection |
| | B | $BD_8 \geq 1$ &<br>$BD_9 < BD_8$ &<br>$BD_{10} < BD_9$ | Seropositive at BD8 followed by waning COI from BD 8 to BD 9 and BD 9 to BD 10 | B | NA | Non-infection |
| | C | $BD_8 \geq 1$ &<br>$BD_9 > BD_8$ &<br>$BD_{10} < BD_9$ | Seropositive at BD8 followed by boosting COI from BD8 to BD9 and waning COI from BD9 to BD10 | C<sub>1</sub> | $BD_9/BD_8 > \gamma$ | Non-infection |
| | | | | C<sub>0</sub> | $BD_9/BD_8 \leq \gamma$ | Non-infection |
| | D | $BD_8 < 1$ &<br>$BD_9 > 1$ &<br>$BD_{10} < BD_9$ | Seronegative at BD8 followed by seroconversion at BD9 then waning COI from BD9 to BD10 | D | NA | Non-infection |
| | E | $BD_8 \geq 1$ &<br>$BD_9 > BD_8$ &<br>$BD_{10} > BD_9$ | Seropositive at BD8 followed by boosting COI from BD8 to BD9 and another boosting COI from BD9 to BD10 | E<sub>2</sub> | $BD_{10}/BD_9 > \gamma$ | Reinfection |
| | | | | E<sub>1</sub> | $BD_{10}/BD_9 \leq \gamma$ &<br>$BD_9/BD_8 > \gamma$ | Non-infection |
| | | | | E<sub>0</sub> | $BD_{10}/BD_9 \leq \gamma$ &<br>$BD_9/BD_8 \leq \gamma$ | Non-infection |
| | F | $BD_8 < 1$ &<br>$BD_9 \geq 1$ &<br>$BD_{10} > BD_9$ | Seronegative at BD8 followed by seroconversion at BD9 and boosting COI from BD9 to BD10 | F<sub>1</sub> | $BD_{10}/BD_9 > \gamma$ | Reinfection<sup>†</sup> |
| | | | | F<sub>0</sub> | $BD_{10}/BD_9 \leq \gamma$ | Non-infection |
| | G | $BD_8 \geq 1$ &<br>$BD_9 < BD_8$ &<br>$BD_{10} > BD_9$ | Seropositive at BD8 followed by waning COI from BD8 to BD9 and boosting COI from BD9 to BD10 | G<sub>1</sub> | $BD_{10}/BD_9 > \gamma$ | Reinfection |
| | | | | G<sub>0</sub> | $BD_{10}/BD_9 \leq \gamma$ | Non-infection |
| | H | $BD_8 < 1$ &<br>$BD_9 < 1$ &<br>$BD_{10} \geq 1$ | Seronegative at BD8 and BD9 followed by seroconversion at BD10 | H | NA | Primary infection or infection<sup>‡</sup> |

<sup>\*</sup>Outcome of whether a specific serologic category is considered as a marker of infection between mid-point (of BD 8 and BD 9), and BD 10.
<sup>#</sup>Refined categorization differentiating strong vs weak boosting signal to account for potential measurement noise.
<sup>†</sup>In rare occasions, this could also be primary infection occurred right before BD 9, with COI yet to reach high level at BD 9 and COI at BD 10 still significantly higher than that of BD 9 even after waning.
<sup>‡</sup>This could be primary infection seroconverted between BD 9 and BD 10 or reinfection between BD 9 and BD 10 with prior infection sero-reverted at BD 8 and BD 9.

**Figure S1:**
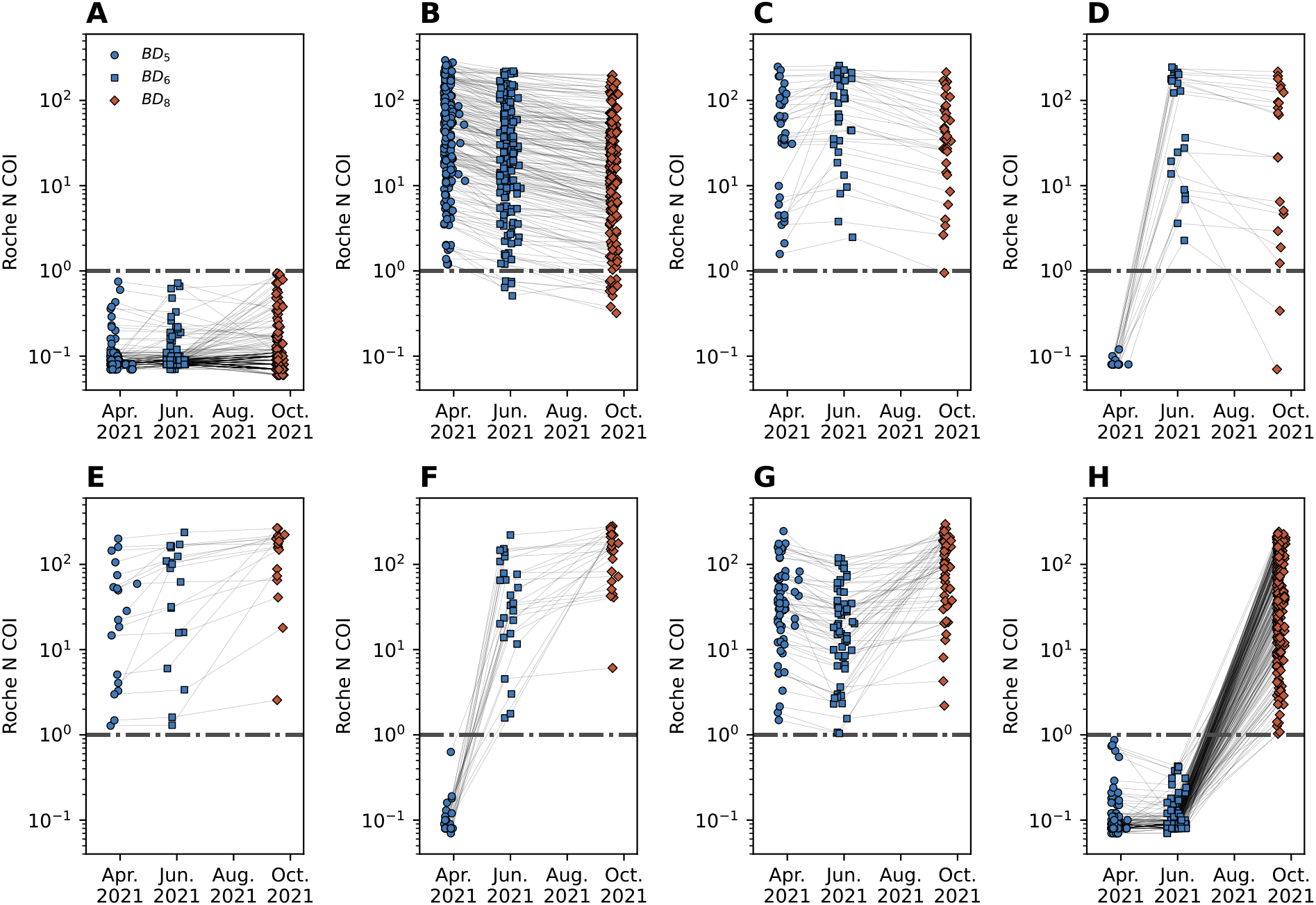
Panel A-H, The serologic trajectory for Roche anti-N COIs at BD 5, BD 6, and BD 8 for each of the crude serologic categories A, B, C, D, E, F, G, and H, respectively. The timing of the BDs bounds SARS-CoV-2 infections during South Africa’s 3^rd^ epidemic wave (Figure 1). The horizontal dashed line indicates the Roche anti-N COI reactivity threshold for seropositivity.

**Figure S2:**
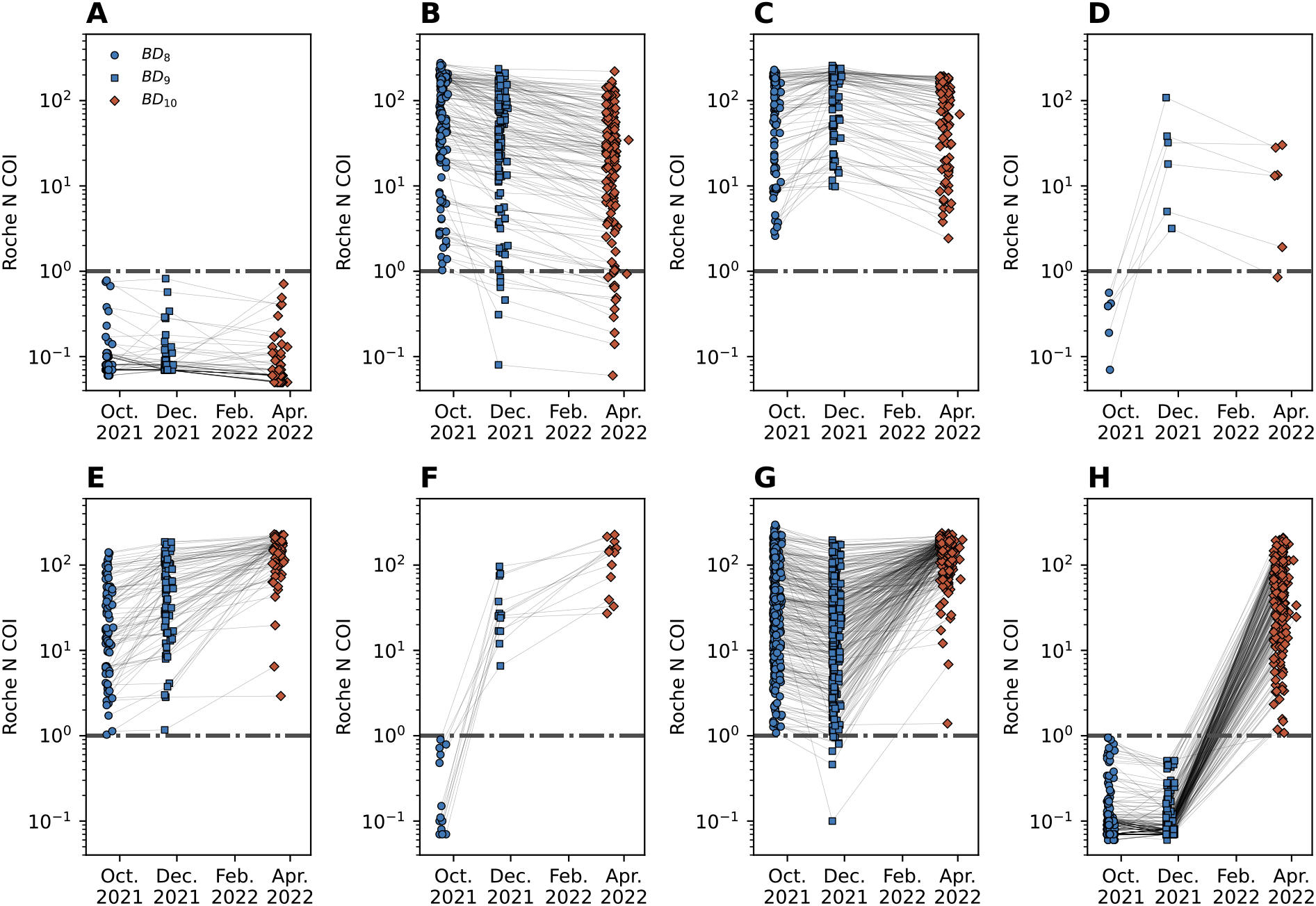
Panel A-H, The serologic trajectory for Roche anti-N COIs at BD 8, BD 9, and BD 10 for each of the crude serologic categories A, B, C, D, E, F, G, and H, respectively. The timing of the BDs bounds SARS-CoV-2 infections during South Africa’s 4^th^ epidemic wave (Figure 1). The horizontal dashed line indicates the Roche anti-N COI reactivity threshold for seropositivity.

**Figure S3:**
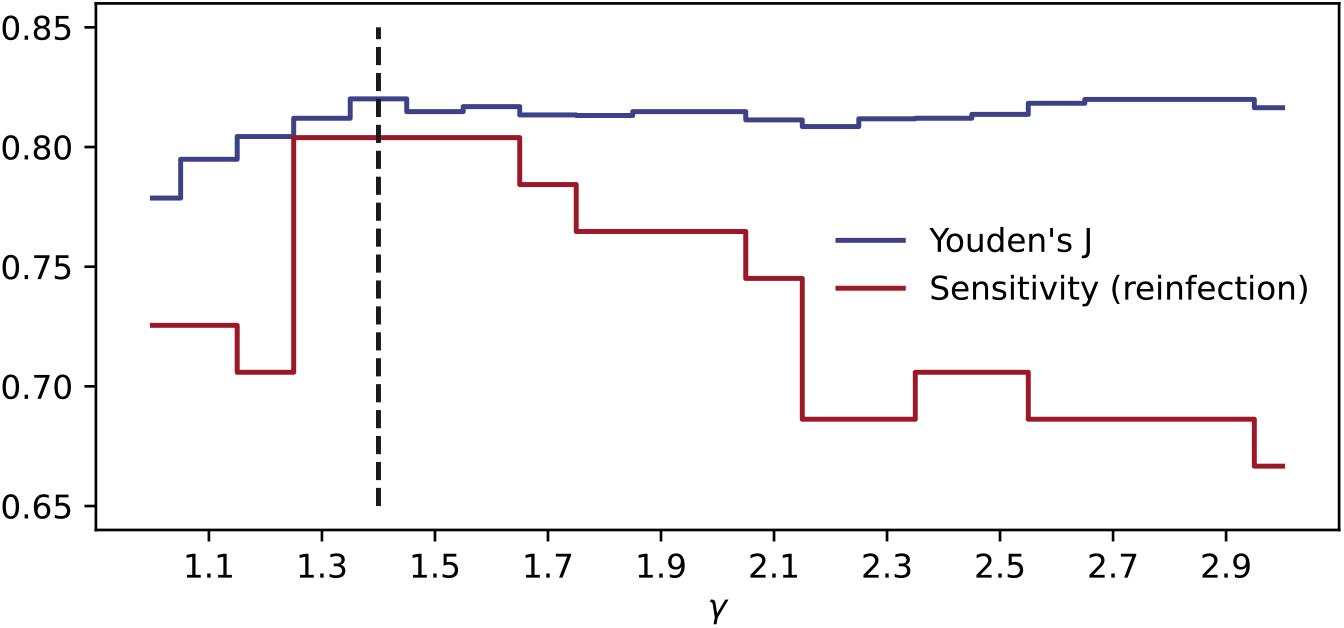
The optimized serial serologic patterns to categorize SARS-CoV-2 infections by Youden’s J statistics (blue line) and the corresponding sensitivity for reinfections (red line).

**Figure S4:**
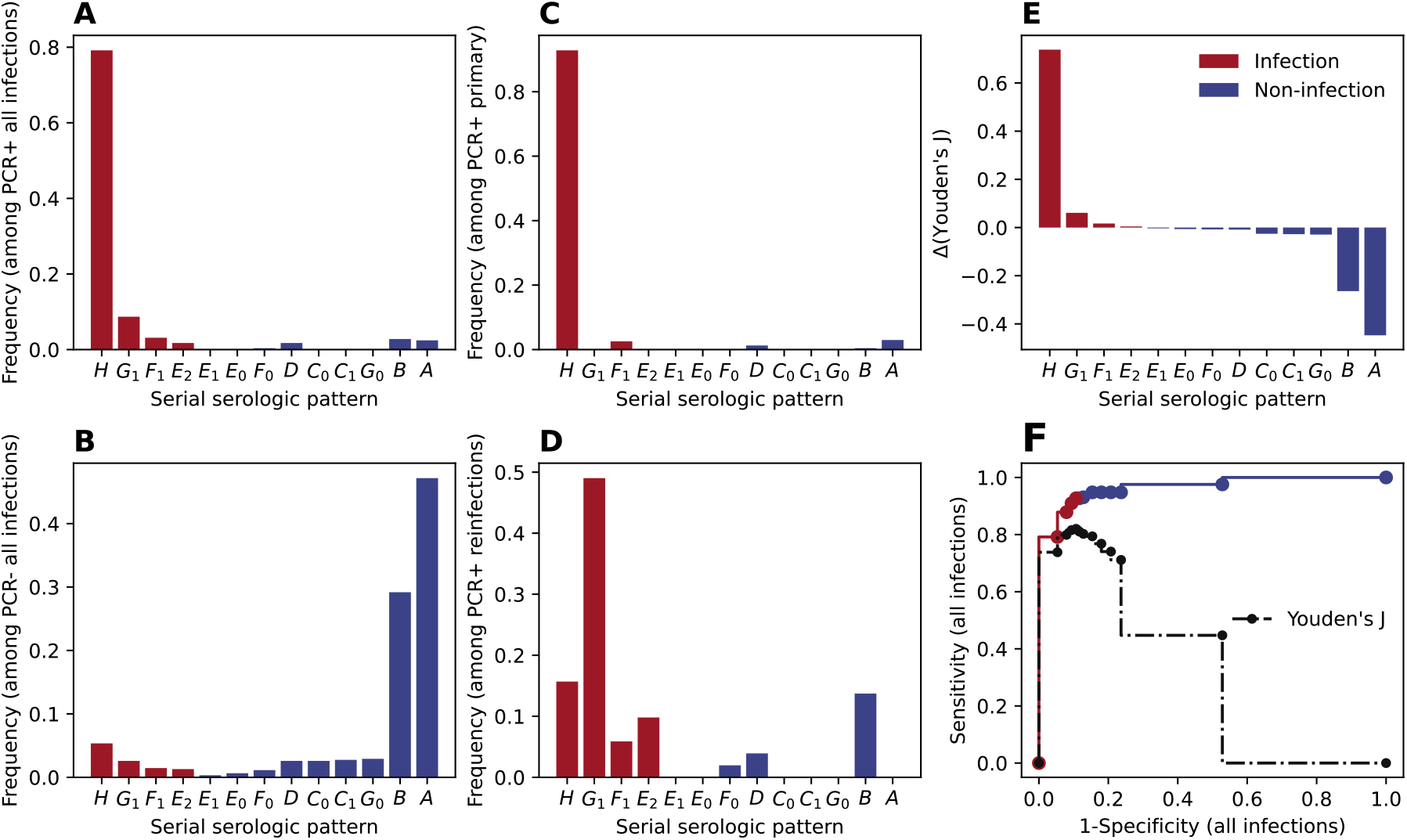
For *γ*=1.4, The optimized serial serologic patterns to categorize SARS-CoV-2 infections by Youden’s J statistics (blue line) and the corresponding sensitivity for reinfections (red line). Panel A: the distribution of serologic patterns among all rRT-PCR confirmed infection (PCR+). B: the distribution of serologic patterns among all rRT-PCR confirmed non-infection (PCR-). C: same as A but among primary infections, D, same as A but among reinfections infections. E, net-contribution of Youden’s J statistic for each of the serial serologic patterns. F, the receiver operating characteristic curve (ROC) by incorporating serial serologic patterns one by one in the order of the net contribution to Youden’s J statistics (panel E); the dashed line is the overall Youden’s J statistics. Across all panels, reds are serial serologic patterns selected as markers for SARS-CoV-2 infection (with net-positive contribution to Youden’s J), while blues are serial serologic patterns selected as marks for SARS-CoV-2 non-infection.

